# Effect of cognitive reserve on physiological measures of cognitive workload in older adults with cognitive impairments

**DOI:** 10.1101/2022.09.08.22279748

**Authors:** Hannes Devos, Kathleen Gustafson, Ke Liao, Pedram Ahmadnezhad, Emily Kuhlmann, Bradley Estes, Laura E. Martin, Jonathan D. Mahnken, William M. Brooks, Jeffrey M. Burns

## Abstract

**Background:** Cognitive reserve may protect against cognitive decline. However, its effect on physiological measures of cognitive workload in adults with cognitive impairments is unclear.

**Objective:** The aim was to determine the association between cognitive reserve and physiological measures of cognitive workload in older adults with and without cognitive impairments.

**Methods:** 29 older adults with cognitive impairment (age: 75±6, 11 (38%) women, MOCA scores 20±7) and 19 with normal cognition (age: 74±6; 11 (58%) women; MOCA 28±2) completed a working memory test of increasing task demand (0-, 1-, 2-back). Cognitive workload was indexed using amplitude and latency of the P3 event-related potential (ERP) at electrode sites Fz, Cz, and Pz, and changes in pupillary size, converted to an index of cognitive activity (ICA). The Cognitive Reserve Index questionnaire (CRIq) evaluated Education, Work Activity, and Leisure Time as a proxy of cognitive reserve.

**Results:** Higher CRIq total scores were associated with larger P3 ERP amplitude (p=0.048), independent of cognitive status (p=0.80), task demand (p=0.003), and electrode site (p<0.0001). This relationship was mainly driven by Work Activity (p=0.0005). Higher CRIq total scores also correlated with higher mean ICA (p = 0.002), regardless of cognitive status (p=0.29) and task demand (p=0.12). Both Work Activity (p=0.0002) and Leisure Time (p=0.045) impacted ICA. No relationship was found between CRIq and P3 latency.

**Conclusion:** Cognitive reserve affects cognitive workload and neural efficiency, regardless of cognitive status. Future longitudinal studies should investigate the causal relationship between cognitive reserve and physiological processes of neural efficiency across cognitive aging.

## INTRODUCTION

Cognitive reserve refers to the adaptability of the brain to cope with the effects of normal and pathological aging on cognitive functioning [1, 2]. Various factors, including advanced education, intellectually stimulating work activities, and a rich social life, influence cognitive reserve [1, 3]. Older adults with greater cognitive reserve experience less age-related cognitive decline and may have a reduced risk of dementia [4-7].

Cognitive reserve may also impact the cognitive workload needed to execute a task. Cognitive workload reflects the exerted mental and physical effort in response to the cognitive demands and time constraints of a task [8]. Cognitive workload increases linearly with task demand until the cognitive resources available to complete the task are depleted. When cognitive overload occurs, task performance will decrease [9]. Older age and age-related neurodegeneration may affect the availability of cognitive resources, resulting in increased cognitive workload to execute the task [9]. Such increased cognitive workload has been observed in older adults with normal cognition and cognitive impairments, even with equal performance on cognitive tasks [9]. The increased cognitive workload observed in both normal and pathological cognitive aging is likely to reflect either decreased neural efficiency or compensatory mechanisms to cope with the demands of the task [9].

Electro-encephalography (EEG) and pupillary recording are two relatively inexpensive and non-intrusive physiological measures with excellent temporal resolution to evaluate cognitive workload in real-time [9]. Event-related potentials (ERP) are very small voltages generated in pyramidal neurons of the cortex in response to specific events or stimuli recorded using EEG [10]. The P3 component is the third positive waveform in the ERP that occurs at about or slightly later than 300 ms after stimulus presentation [11]. The P3 is believed to be associated with cognitive resource allocation during information processing, memory encoding, and updating of information [12, 13]. Working memory tasks are widely used paradigms in P3 ERP studies across the spectrum of cognitive impairments, as deterioration in working memory is one of the earliest cognitive dysfunctions observed in mild cognitive impairment (MCI) and is a reliable predictor of Alzheimer’s disease (AD) [14, 15]. Although P3 has been used in healthy populations to explain the neural substrates of cognitive reserve, very few studies have investigated the relationship between cognitive reserve and P3 in clinical populations [16]. Whereas higher cognitive reserve affects P3 amplitude and latency with respect to task demand in healthy older adults, no such relationship has been observed in MCI [17]. Changes in pupillary size have also shown to correlate with cognitive workload in older adults with and without cognitive impairments,[18-20] but no studies have evaluated the effect of cognitive reserve on pupillary response in people with cognitive impairments.

As cognitive reserve plays a crucial role in slowing down progression of MCI and AD, the aim of this study was to elucidate the effect of cognitive reserve on physiological measures of cognitive workload. We hypothesized that higher cognitive reserve will be associated lower cognitive workload, indexed by larger P3 amplitudes (reflective of neural efficiency) and lower pupillary response, independent of cognitive status and task demand.

## MATERIALS AND METHODS

### Participants

Participants were recruited between 5/30/2018 and 02/25/2022 from the University of Kansas Alzheimer’s Disease Research Center (KU ADRC). Inclusion criteria were: (1) age 65 years or older; (2) understanding of all instructions in English; or (3) informed consent. Exclusion criteria were: (1) currently taking steroids, benzodiazepines, or neuroleptics; (2) history of any substance abuse; or (3) any contra-indications to EEG.

The Clinical Dementia Rating [21] and the Uniform Data Set neuropsychological battery [22] were administered to determine cognitive status (normal or impaired). The team administering the cognitive tests, EEG, and pupillary recordings were blind to cognitive status of participants.

Participants with normal cognition had previously undergone a PET scan of the brain to rule out increased amyloid-β depositions. The protocol has been published elsewhere [23]. Three raters interpreted the PET scans to rule out elevated amyloid-β [25, 26].

Participants with cognitive impairments had either mild or major neurocognitive disorder. Seventeen (59%) were categorized as mild neurocognitive disorder (MCI due to probable AD, n = 8; MCI of unknown or mixed etiology, n = 7; vascular MCI, n = 1; and MCI due to fronto-temporal dementia, n = 1). Twelve (41%) were categorized as major neurocognitive disorders (AD, n = 6; dementia of unknown or mixed etiology, n = 4, and Lewy Body Dementia, n = 2).

### Procedure

#### Demographic and Clinical Information

The demographic survey included age, sex, and hand dominance. All participants were right-handed. We screened for cognitive impairments using the Montreal Cognitive Assessment (MOCA) [27].

#### Cognitive Reserve Index Questionnaire

The Cognitive Reserve Index questionnaire (CRIq) is a widely validated questionnaire of cognitive reserve [28]. The CRIq is composed of 20 items, which are categorized in three sections: Education, Working Activity; and Leisure Time. Total CRIq scores are calculated as the average of the section scores, each standardized and transposed to a mean = 100 and standard deviation of 15. Scores lower or equal to 70 are considered low; between 70 and 84 medium-low; between 85 and 114 medium; between 115 and 130 medium-high; and higher than 130 high.

#### N-back Test

The n-back test involved participants pressing a mouse button with the right hand when a white letter that appeared on a black screen was the same as the letter presented n- places back [29, 30]. Participants completed the 0-, 1-, and 2-back test. The n-back tests reflects memory, but higher levels of task demand also assess higher order cognitive functions such as updating of information and maintaining representations of recently presented stimuli [31].

Participants familiarized with each test by practicing a random sequence of 3 targets and 7 nontargets until they felt comfortable to proceed with the actual test. Each n-back test consisted of 60 trials (target, 33.3%) that required a mouse click and 120 trials (nontarget, 66.7%) that did not require a response. All trials were presented in random order. Stimulus onset asynchrony was 2200 ms (presentation time of 500 ms and blank interstimulus interval of 1700 ms). We set a random jitter of +/-50 ms. The total duration of each n-back test was about 400 seconds. The main performance outcome measures were accuracy, defined as the number of correct responses to the 60 trials, and response time in seconds.

#### P3 Event-Related Potential

While completing the n-back tests, participants were fitted with a high-density 256-electrode Geodesic Sensor Net from Magstim EGI. The Net Amps 400 amplifier was used with a bandwidth from DC to 2,000 Hz and input impedance larger than 1 GΩ. The electrode impedance was kept less than either 50 KΩ or 100 KΩ. Continuous EEG was digitized at 1,000 Hz and referenced to Cz without an online filter. All other EEG processes were done in EEGLab [32] and in ERPLab [33]. In the offline process, EEG data were band-pass filtered between 0.5 Hz and 30 Hz in Magstim EGI NetStation Tools software with a roll-off frequency of 0.3 Hz. Electrodes around the face were removed from data processing, leaving 183 channels to be analyzed. Bad channels were removed through visual inspection of data and automatic identification using joint probability methods with a z-score kurtosis cut-off of 6. Subsequently, independent component analysis was employed to detect and remove various ocular and movement artifacts or cardiovascular signals. Continuous EEG data were segmented into stimulus-locked ERP’s with an epoch interval of 100 ms before to 1000 ms after stimulus onset. Epochs were baseline corrected using the prestimulus interval. Epochs of incorrect and missed responses were manually removed. Signals from bad electrodes were interpolated using surrounding electrode data. All channels were off-line referenced to the linked mastoid average. ERP data containing more than 50% artifacts (which included noisy epochs and epochs of missed and incorrect responses) were not included in the analyses. Artifacts ranged from 0% to 26% (mean 9% ± 7) of epochs for the 0-back test, 0% to 36% (mean 7% ± 8) for the 1-back test, and 0% to 43% (mean 10% ± 12) for the 2-back test. Thus, no participant was excluded from the ERP analyses. The measurement window of the P3 ERP was established *a priori* and ranged between 250 ms and 650 ms, based on our preliminary data [34]. We identified a priori Fz as the main electrode site because of the prefrontal cortex involvement in working memory, and the shift from parietal to frontal brain areas in older adults [35]. However, we also report P3 ERP from Cz and Pz. The P3 amplitude and latency at these electrode sites were calculated as the average of the clusters of surrounding channels [36].

Since the P3 difference waveform included both a negative and positive component (Figure 1), we calculated the rectified area amplitude within the P3 time window of the task effect (target – nontarget). P3 latency was calculated as the 50% fractional area latency, which is the midpoint of the component that divided that area under the curve in two equal regions. Fractional area latency is typically recommended for difference waves of P3 components [37].

**Figure 1.**
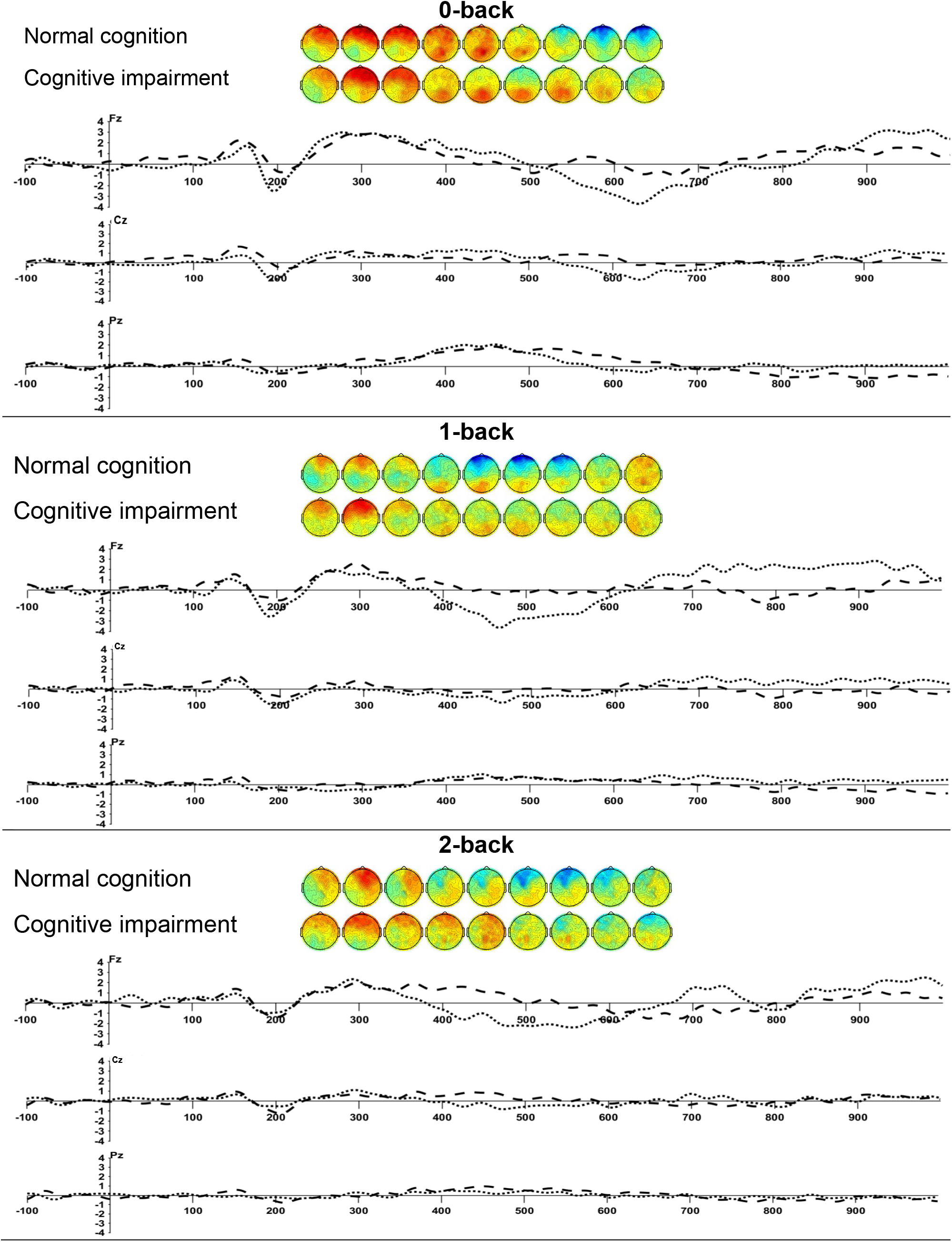
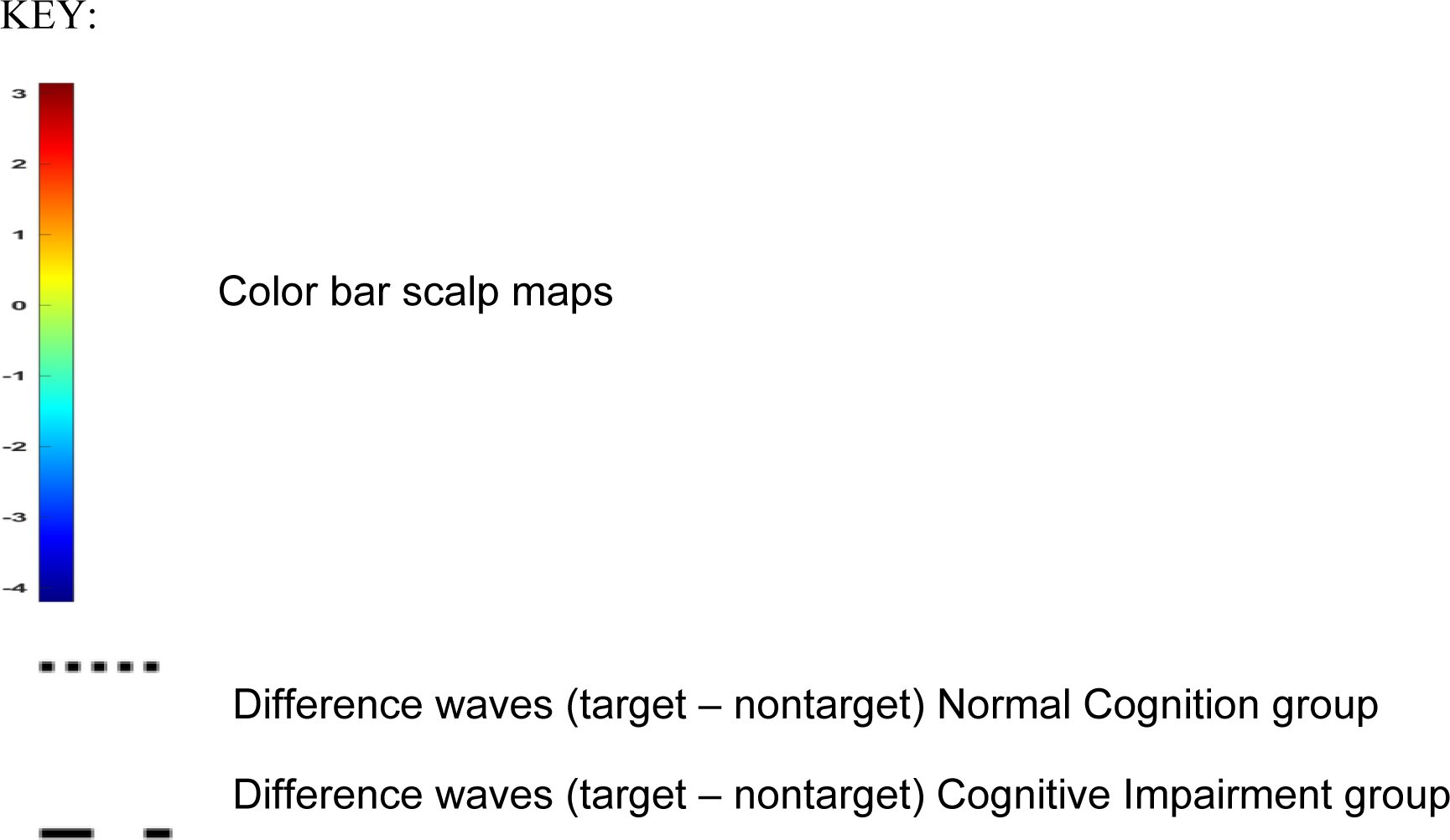
Scalp maps (from 250 to 650 ms in 50 ms increments) and difference waves (target – nontarget) of the P3 event-related potential of the 0-back, 1-back, and 2-back tests in the groups with normal cognition (n=19) and cognitive impairments (n = 29)

#### Index of Cognitive Activity

A remote infrared eye tracker (FX3, SeeingMachines, Inc, Canberra, Australia) was mounted right below the computer screen to record pupillary data of both eyes while completing the n-back tests. Pupillary data were recorded at 60 Hz using EyeWorks Record and analyzed using Eyeworks Analyze (Eye Tracking, Inc, Solana Beach, CA). We required participants to focus on the screen to minimize potential artifacts of the light reflex and eye movements on pupillary recording. However, the pupil continues to oscillate irregularly even when controlling for ambient lighting. The Index of Cognitive Activity (ICA) decomposes the raw pupillary size to different wavelets of high and low frequency components of the signal [38]. Doing so, changes in pupillary size in response to cognitive workload are separated from changes in pupillary size due to the light reflex [8, 39]. The ICA is calculated by dividing the number of rapid small pupillary dilations per second by the number of expected rapid pupillary dilations per second [38]. The values are then transformed using the hyperbolic tangent function. Blinks are factored out by linear interpolation of adjacent time spans to produce continuous values ranging between 0 and 1 [38]. Missing data ranged from 0 – 55% (mean 3 ± 8%) for the left eye and 0 to 96% (mean 4 ± 14%) in the right eye. Since the left eye produced more accurate recordings, we only reported mean ICA of the left eye in the results. No participants were excluded from the analyses because of excess (>50%) missing data.

### Data Analysis

Unpaired t-tests, Wilcoxon Rank Sum tests, and Chi-square tests were used to compare differences in descriptive and behavioral variables between groups. Linear mixed models were employed to identify the relationship between CRIq and physiological outcomes. We used a random intercept term with a subject-specific coefficient to adjust for correlation between measures within subjects. For P3 amplitude and latency, we entered two main within-group effects of task demand (0-, 1-, 2-back) and electrode sites (Fz, Cz, Pz), and one main between-group effect of cognitive status (normal cognition, cognitive impairment). For ICA, we used one main within-group effect (task demand) and one main between-group effect (cognitive status). We entered CRIq scores as covariate and cognitive status*task demand as interaction effect into the model. Post-hoc Sidak correction was applied for pairwise comparisons. Residuals of all outcome variables were normally distributed. We adjusted for potential confounders such as age and sex in separate linear mixed models. P values of 0.05 or less were considered significant. All analyses were performed in SAS, version 9.4.

## RESULTS

### Participant Characteristics

We consented 29 older adults with cognitive impairments (age: 74 ± 6, 11 (38%) women, MOCA 20 ± 7) and 19 with normal cognition (age: 74 ± 6; 11 (58%) women; MOCA 28 ± 2). Participants with cognitive impairments scored lower on the MOCA and on all behavioral outcomes, except for accuracy on the 0-back test (**Table 1**). No differences were found in CRIq total or item scores between groups. CRIq scores ranged from 81 to 158 in the normal cognition group: one participant (5%) scored low; 3 (16%) scored medium-low; 9 (47%) scored medium-high; and 6 (32%) scored high. Scores in the group with cognitive impairments ranged from 104 to 146: nine (32%) scored medium; 10 (34%) scored medium-high; and 10 (34%) scored high.

**Table 1.**
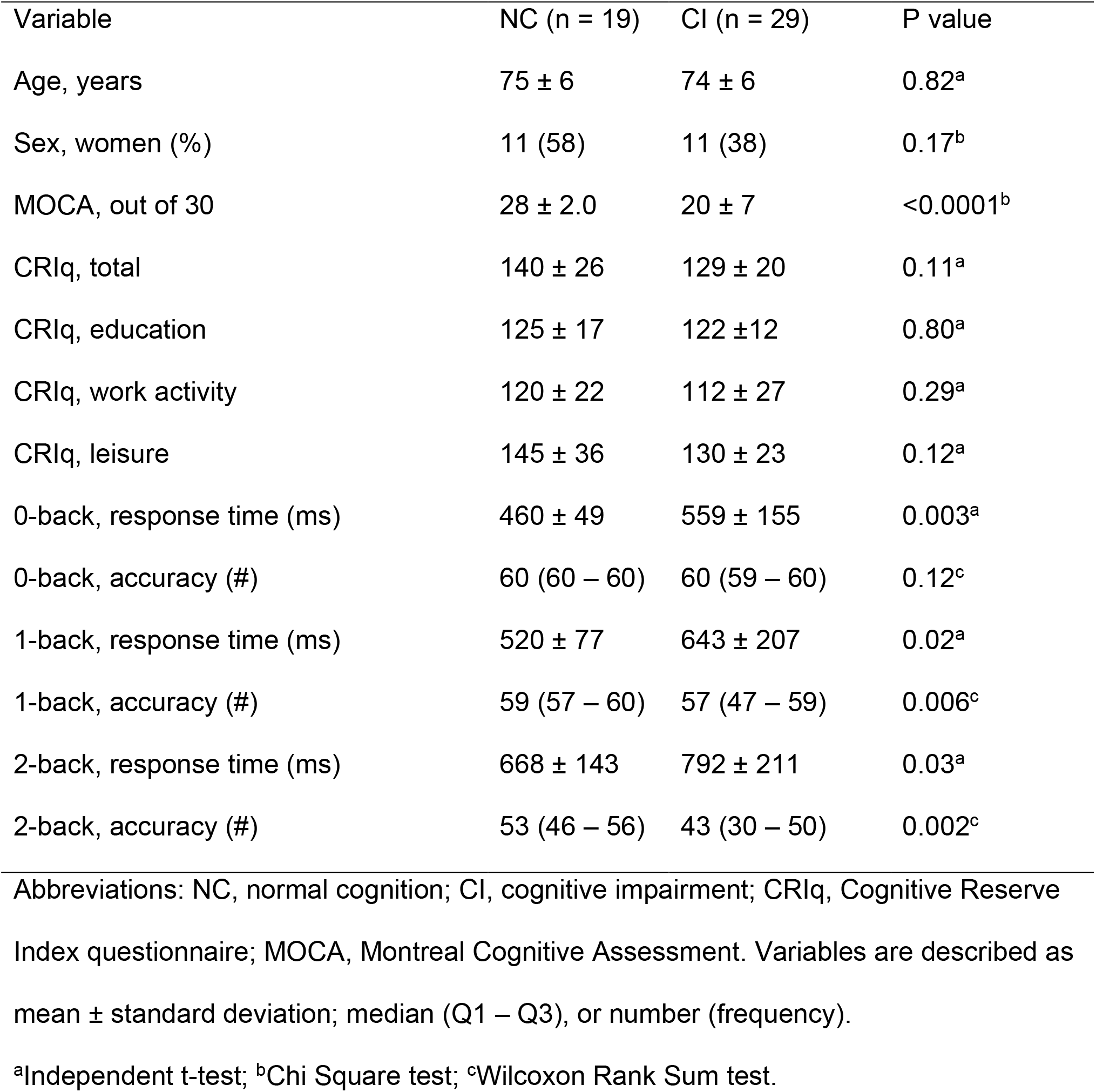
Comparison of clinical and behavioral variables between older adults with normal cognition (NC) and cognitive impairments (CI)

| Variable | NC (n = 19) | CI (n = 29) | P value |
| --- | --- | --- | --- |
| Age, years | 75 ± 6 | 74 ± 6 | 0.82<sup>a</sup> |
| Sex, women (%) | 11 (58) | 11 (38) | 0.17<sup>b</sup> |
| MOCA, out of 30 | 28 ± 2.0 | 20 ± 7 | <0.0001<sup>b</sup> |
| CRIq, total | 140 ± 26 | 129 ± 20 | 0.11<sup>a</sup> |
| CRIq, education | 125 ± 17 | 122 ± 12 | 0.80<sup>a</sup> |
| CRIq, work activity | 120 ± 22 | 112 ± 27 | 0.29<sup>a</sup> |
| CRIq, leisure | 145 ± 36 | 130 ± 23 | 0.12<sup>a</sup> |
| 0-back, response time (ms) | 460 ± 49 | 559 ± 155 | 0.003<sup>a</sup> |
| 0-back, accuracy (#) | 60 (60 – 60) | 60 (59 – 60) | 0.12<sup>c</sup> |
| 1-back, response time (ms) | 520 ± 77 | 643 ± 207 | 0.02<sup>a</sup> |
| 1-back, accuracy (#) | 59 (57 – 60) | 57 (47 – 59) | 0.006<sup>c</sup> |
| 2-back, response time (ms) | 668 ± 143 | 792 ± 211 | 0.03<sup>a</sup> |
| 2-back, accuracy (#) | 53 (46 – 56) | 43 (30 – 50) | 0.002<sup>c</sup> |

Abbreviations: NC, normal cognition; CI, cognitive impairment; CRIq, Cognitive Reserve Index questionnaire; MOCA, Montreal Cognitive Assessment. Variables are described as mean ± standard deviation; median (Q1 – Q3), or number (frequency).
<sup>a</sup>Independent t-test; <sup>b</sup>Chi Square test; <sup>c</sup>Wilcoxon Rank Sum test.

### P3 Event-Related Potential Waveforms

The grand average waveforms of the targets and non-targets at electrode sites Fz, Cz, and Pz for each n-back test are shown in Supplementary Figure 1. The task effect, depicted as the difference wave and scalp maps of targets – nontargets of the P3, between the normal cognition and cognitive impairment groups at the three channels for each n-back test is shown in Figure 1.

Outcomes of the physiological variables are described in Table 2.

**Table 2.**
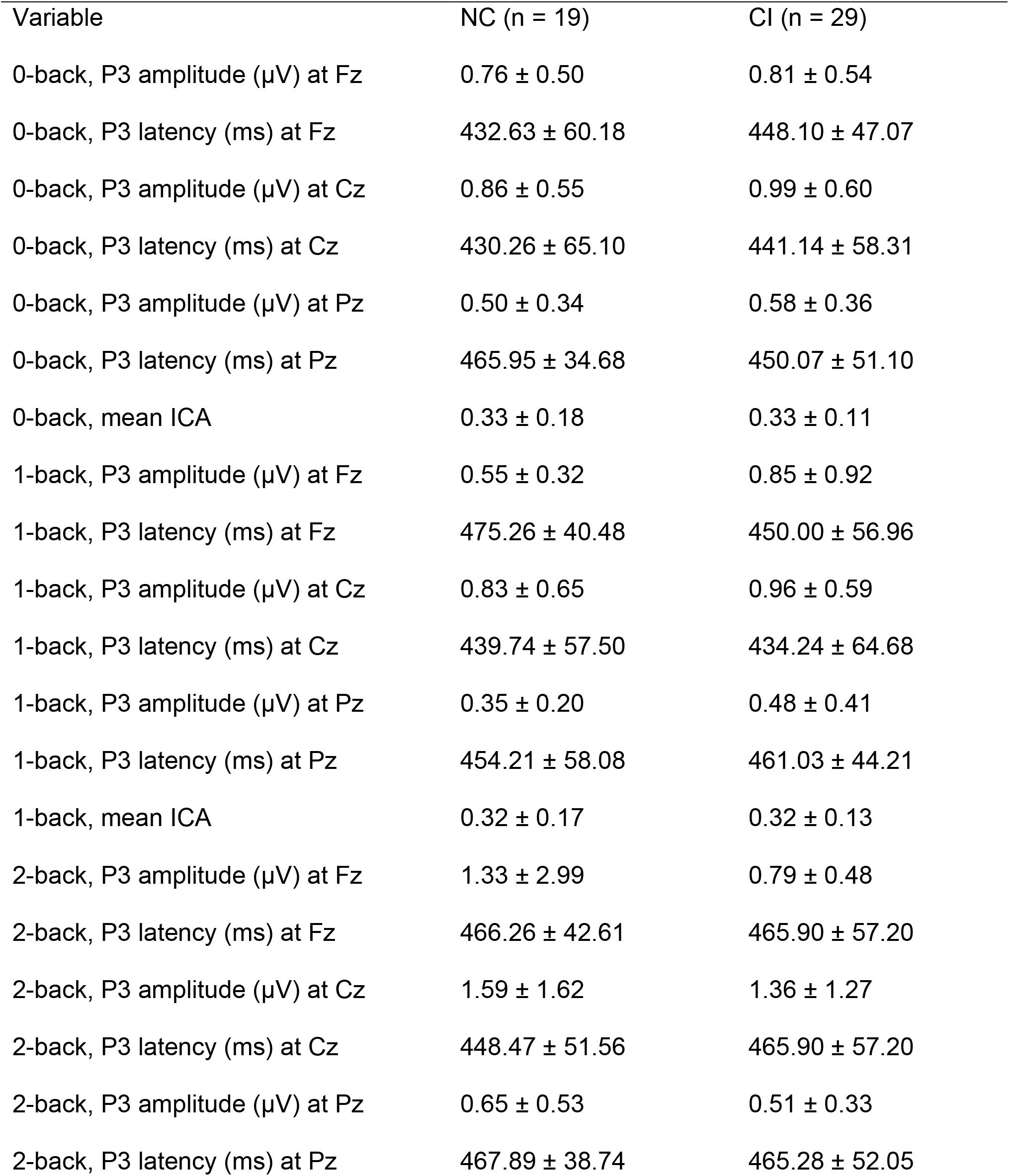
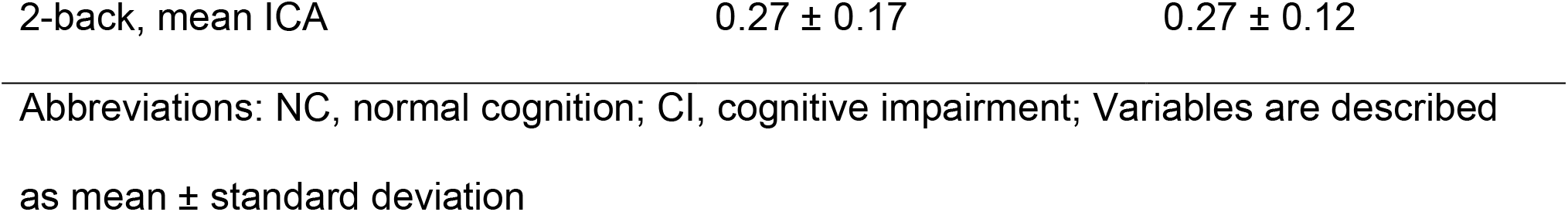
Comparison of physiological variables between older adults with normal cognition and cognitive impairments for each n-back test

| Variable | NC (n = 19) | CI (n = 29) |
| --- | --- | --- |
| 0-back, P3 amplitude (μV) at Fz | 0.76 ± 0.50 | 0.81 ± 0.54 |
| 0-back, P3 latency (ms) at Fz | 432.63 ± 60.18 | 448.10 ± 47.07 |
| 0-back, P3 amplitude (μV) at Cz | 0.86 ± 0.55 | 0.99 ± 0.60 |
| 0-back, P3 latency (ms) at Cz | 430.26 ± 65.10 | 441.14 ± 58.31 |
| 0-back, P3 amplitude (μV) at Pz | 0.50 ± 0.34 | 0.58 ± 0.36 |
| 0-back, P3 latency (ms) at Pz | 465.95 ± 34.68 | 450.07 ± 51.10 |
| 0-back, mean ICA | 0.33 ± 0.18 | 0.33 ± 0.11 |
| 1-back, P3 amplitude (μV) at Fz | 0.55 ± 0.32 | 0.85 ± 0.92 |
| 1-back, P3 latency (ms) at Fz | 475.26 ± 40.48 | 450.00 ± 56.96 |
| 1-back, P3 amplitude (μV) at Cz | 0.83 ± 0.65 | 0.96 ± 0.59 |
| 1-back, P3 latency (ms) at Cz | 439.74 ± 57.50 | 434.24 ± 64.68 |
| 1-back, P3 amplitude (μV) at Pz | 0.35 ± 0.20 | 0.48 ± 0.41 |
| 1-back, P3 latency (ms) at Pz | 454.21 ± 58.08 | 461.03 ± 44.21 |
| 1-back, mean ICA | 0.32 ± 0.17 | 0.32 ± 0.13 |
| 2-back, P3 amplitude (μV) at Fz | 1.33 ± 2.99 | 0.79 ± 0.48 |
| 2-back, P3 latency (ms) at Fz | 466.26 ± 42.61 | 465.90 ± 57.20 |
| 2-back, P3 amplitude (μV) at Cz | 1.59 ± 1.62 | 1.36 ± 1.27 |
| 2-back, P3 latency (ms) at Cz | 448.47 ± 51.56 | 465.90 ± 57.20 |
| 2-back, P3 amplitude (μV) at Pz | 0.65 ± 0.53 | 0.51 ± 0.33 |
| 2-back, P3 latency (ms) at Pz | 467.89 ± 38.74 | 465.28 ± 52.05 |
| 2-back, mean ICA | $0.27 \pm 0.17$ | $0.27 \pm 0.12$ |

Abbreviations: NC, normal cognition; CI, cognitive impairment; Variables are described as mean ± standard deviation

### P3 amplitude

The linear mixed model unadjusted for age and sex showed a significant effect of total CRIq scores (F = 3.92; p = 0.048), task demand (6.06; p = 0.003), electrode site (F = 15.78; p < 0.0001) and interaction effect of cognitive status*task demand (F = 3.17; p = 0.04) on P3 amplitude. No main effects of cognitive status on P3 amplitude were found (F = 0.007; p = 0.80). Post-hoc analyses demonstrated that higher total CRIq scores were associated with larger P3 amplitude (β = 0.004 ± (standard error of measurement) 0.002; t = 1.98; p = 0.048). Although the main effect of task demand was associated with lower P3 amplitude, post-hoc analyses corrected for multiple comparisons did not reach significance. The post-hoc analysis of the interaction effect of cognitive status*task demand demonstrated that the P3 amplitude of the 0-back test (β = −0.48 ± 0.14; t = −1.94; p = 0.05) and the 1-back test (β = −0.34 ± 0.14; t = −2.36; p = 0.02) was lower compared to the 2-back, but only in the group with normal cognition. No such effects were observed in the group with cognitive impairments. P3 amplitude was larger at Fz (β = 0.17 ± 0.07; t = 2.12; p = 0.04) and at Cz (β = 0.39 ± 0.07; t = 5.49; p<0.0001) compared to Pz.

When adjusted for age (F = 1.79; p = 0.18) and sex (F = 0.27; p = 0.59), the variables task demand (F = 6.06; p = 0.003), cognitive status*task demand (F = 3.17; p = 0.04), and electrode site (F = 15.78; p<0.0001) remained significant in the model. In contrast, cognitive status (F = 0.14; p = 0.71) and CRIq scores (F = 2.53; p = 0.11) were no longer significant.

CRIq item scores were also entered in separate linear mixed models. Education (unadjusted, F = 3.55; p = 0.06; adjusted, F = 1.74; p = 0.19) and Leisure Time (F = 2.83; p = 0.06; adjusted F = 0.15; p = 0.70) were not significant, whereas higher Work Activity (unadjusted, F = 12.33; p = 0.0005; adjusted, F = 9.21; p = 0.003) was associated with increased P3 amplitude.

### P3 latency

Task demand (F = 3.50; p = 0.03) and electrode sites (F = 5.180; p = 0.007) were the only variables significantly associated with P3 latency in the unadjusted model. Latency was shorter in the 0-back compared to the 2-back (−16.67 ± 8.19; t = −1.97; p = 0.049). P3 latency was significantly shorter at Cz compared to Pz (−19.22 ± 6.32; t = −3.04; p = 0.003). CRIq scores (F = 0.50; p = 0.48), cognitive status (F = 0.02; p = 0.89) and cognitive status*task demand (F-0.35; p = 0.71) were not significantly correlated with P3 latency.

When adjusting the model for age (F = 4.12; p = 0.04) and sex (F = 0.39; p = 0.53), task demand (F = 3.53; p = 0.03) and electrode site (F = 5.13; p = 0.006) remained significant in the model.

### Index of Cognitive Activity

Whereas significant effects were found of CRIq scores (F = 10.54; p = 0.002) on mean ICA, no such effects were found of cognitive status (F = 1.13; p = 0.29), task demand (F = 2.13; p = 0.12) and cognitive status*task demand (F = 0.01; p = 0.99). Higher CRIq scores were associated with lower mean ICA scores (−0.002 ± 0.0006; t = −3.25; p = 0.002).

When adjusted for age (F = 1.34; p = 0.25) and sex (F = 0.07; p = 0.80), CRIq scores remained significant (F = 8.30; p = 0.005). Education (unadjusted, F = 0.16; p = 0.69; adjusted F = 0.00; p = 0.97) did not impact ICA. Work Activity (unadjusted, F = 14.66; p = 0.0002; adjusted, F = 12.26; p = 0.0006) and Leisure Time (unadjusted, F = 4.09; p = 0.045; adjusted, F = 3.77; p = 0.05) were significantly associated with ICA.

## DISCUSSION

The aim of this study was to evaluate the association between cognitive reserve and physiological measures of cognitive workload in older adults with and without cognitive impairments. Higher cognitive reserve was associated with increased P3 ERP amplitude and lower ICA, independent of cognitive status and task demand. In particular, Work Activity and Leisure of the CRiq showed the greatest associations with physiological measures of cognitive workload.

Older adults with higher cognitive reserve showed larger P3 amplitude, reflecting more efficient post-stimulus categorization processing [40]. This relationship manifested independent of cognitive status. Therefore, cognitive reserve directly impacts the efficiency of neural processing, supporting previous research [17]. However, Gu et al found distinct mediation processes of cognitive reserve and P3 ERP in older adults without and with cognitive impairments [17]. Whereas higher cognitive reserve affected efficiency of neural processing in normal cognitive aging, no such effects were found in patients with amnestic MCI [17]. Our results indicate that cognitive reserve continues to play a protective role in efficiency of neural processing, even in individuals with marked cognitive decline.

Previous studies using functional magnetic resonance imaging (fMRI) corroborate the protective mechanisms of cognitive reserve across the spectrum of cognitive aging [41]. Higher cognitive reserve was related to increased frontal activity in older adults with cognitive impairments compared to those with normal cognition [42]. The compensatory processes observed in the frontal areas may likely lead to more efficient neural processing in individuals with cognitive impairments who have higher cognitive reserve. The higher rectified area P3 amplitudes in the central and frontal channels compared to the parietal channels support this hypothesis. However, the time window for modulation of cognitive reserve on brain reserve may be limited to individuals with MCI [43]. Higher cognitive reserve in older adults with amnestic MCI was associated with lower macromolecular tissue volume across major white tracts, whereas no such relationship was observed in individuals with AD [44]. Likewise, older adults with MCI and high cognitive reserve manifested decreased cortical thickness in the right temporal and in the left prefrontal lobe, and increased fractional dimension in the right temporal and in the left temporo-parietal lobes compared to those with lower reserve [43]. Future studies combining the spatial accuracy of MRI and the temporal dynamics of EEG may elucidate the mediating effects of cognitive reserve between localized brain compensatory activity and neural efficiency across the cognitive aging spectrum.

Cognitive reserve was also associated with the ICA, a measure of cognitive workload based on moment-to-moment changes in pupillary size [38]. Pupillary dilation in response to cognitive workload is mainly mediated through the locus coeruleus [45]. The locus coeruleus is the main supplier of noradrenaline in the brain and critical in the regulation of physiological arousal and cognition [46]. In vivo and post-mortem imaging studies demonstrate that older adults with cognitive impairments exhibit decreased neuronal density and early tau accumulation in the locus coeruleus [47]. These neurobiological abnormalities in the locus coeruleus result in increased pupillary dilation during cognitive tasks, even in the absence of impairments in cognitive performance [20, 48]. The noradrenergic theory of cognitive reserve postulates that continuous upregulation of noradrenaline in the brain throughout the lifespan is paramount to building cognitive reserve [49]. Individuals who continuously engage in cognitively stimulating activities in their lifetime may exhibit an overall higher noradrenergic tone, building more resilient neurobiological networks that protect against age-related neurodegeneration. Our results support previous studies identifying the role of the locus coeruleus noradrenergic system as a key mediator of cognitive reserve [50].

Work activity and leisure time emerged as the most important cognitive stimulating life events that correlated with physiological measures of cognitive workload. Whereas education had a moderating effect on the relationship between hypometabolism and cognition, previous work activities had a moderating effect on the relationship between cortical atrophy and cognition in AD [51]. Our results are particularly encouraging as occupational attainment and participation in leisure activities are modifiable lifestyle choices that may directly impact cognitive workload.

Our study has several limitations. First, our study included participants with different etiologies and degrees of cognitive impairment, making it difficult to pinpoint the neurobiological mechanisms underlying the link between cognitive reserve and physiological measures of cognitive workload. However, our results also demonstrate a strong association between cognitive reserve and physiological measures of cognitive workload, regardless of disease pathology. Our methods could therefore be applied to different types of dementia. Second, our participants were recruited from a volunteer registry database. This selection bias shows in the relatively high average CRIq scores. Caution is therefore warranted extrapolating our results to the population of older adults with and without cognitive impairments. Third, our ERP measures lacked spatial resolution. Future combined fMRI/EEG studies may elucidate novel mechanisms on the temporal and spatial dynamics of cognitive reserve in older adults across the spectrum of cognitive aging. Fourth, the cross-sectional design of our study prevents making any inferences about the causal relationships between cognitive reserve and physiological measures of cognitive workload. Finally, our results need to be validated in cognitive domains other than working memory.

## Conclusions

Cognitive reserve directly affects physiological measures of cognitive workload across the cognitive aging spectrum. Higher cognitive reserve is associated with more efficient neural processing and decreased cognitive workload to accomplish working memory tasks. Furthermore, our results suggest occupational attainment and participation in leisure activities are modifiable lifestyle choices that may directly benefit cognitive reserve. Future longitudinal studies should investigate the causal relationship between cognitive reserve and physiological processes of neural efficiency in normal and pathological cognitive aging.

## Data Availability

All data produced in the present study are available upon reasonable request to the authors

## Acknowledgments

This work was completed at the Hoglund Biomedical Imaging Center which is supported by the Forrest and Sally Hoglund and a High-End Instrumentation grant from the National Institutes of Health (S10 RR29577). The authors thank all participants for volunteering their time. Research reported in this publication was supported by the National Institute on Aging of the National Institutes of Health under Award Number K01 K01 AG058785 and P30 AG072973. This study was supported in part by a pilot grant of the KU Alzheimer Disease Research Center (P30 AG035982). The content is solely the responsibility of the authors and does not necessarily represent the official views of the National Institutes of Health.

## Availability of Data and Materials

The datasets used and/or analyzed during the current study are available from the corresponding author on reasonable request.

## DECLARATIONS

### Ethics Approval and Consent to Participate

This study was approved by the Human Subjects Committee the University of Kansas Medical Center (#4461). Informed consent was obtained from all participants.

**Consent to publication** obtained from that person, or in the case of children, their parent or legal guardian. All presentations of case reports must have consent for publication.

Not applicable.

## Authors’ contributions

HD, KG, WB, JM, and JB conceptualized the study. HD, KL, PA administered the EEG assessments. HD, KG, and KL created the pipeline for EEG processing. HD, BE, EK processed the EEG data. HD and JM created the statistical plan and conducted the statistical analyses. HD, KG, KL, WB, JM, JB, PA, LM, EK, BE interpreted the results. HD wrote the first version of the manuscript. KG, KL, WB, JM, JB, PA, LM, BE, EK reviewed and revised the manuscript. All authors read and approved the final manuscript.

